# Assessment on Uptake of Self-Care Services among Adolescents and Young People Living With HIV: A Case of Four Facilities in Kitwe District, Zambia

**DOI:** 10.64898/2026.02.02.26345430

**Authors:** Elvis Dala Makukula, Samson Shumba, Chishimba Nakamba Mulambia, Choolwe Jacobs

## Abstract

Adolescents and young people living with HIV (AYPLHIV) face persistent challenges related to stigma, autonomy, and sustained engagement in care. Self-care services including tools and practices that enable individuals to manage their health play a critical role in HIV prevention and treatment by promoting adherence, empowerment, quality of life, and improved health outcomes. Despite their importance, evidence on the uptake and determinants of self-care services among AYPLHIV in Zambia remains limited. This study assessed the level of self-care service utilization and associated factors among adolescents and young people receiving HIV care in selected health facilities in Kitwe District.

A facility-based cross-sectional study was conducted between September 2024 and April 2025 among 485 adolescents and young people aged 15–24 years attending four health facilities in Kitwe. Data were collected using structured, pre-tested questionnaires administered by trained research assistants. Data were analysed using Stata version 17. Overall, uptake of self-care services was suboptimal. Slightly more than half of participants (58%) reported ever using self-care services, while a substantial proportion (42%) had never utilized them. Awareness was generally low, with over half (56%) reporting no knowledge of available self-care interventions. Multivariable analysis showed that adolescents with secondary education had significantly lower odds of utilizing self-care services compared to those with no formal education (AOR = 0.54, p = 0.048). Accessibility was positively associated with utilization (AOR = 1.67, p = 0.031), whereas moderate and high levels of stigmatization were strongly associated with reduced uptake (AOR = 0.53, p = 0.022; AOR = 0.63, p = 0.029). Age, gender, peer influence, and provider support were not significantly associated with self-care service utilization.

In conclusion, self-care service uptake among AYPLHIV in Kitwe District remains low, largely driven by limited awareness, accessibility barriers, and persistent stigma. Strengthening awareness campaigns, improving service availability, and implementing stigma-reduction strategies are essential to enhance self-care engagement and optimize HIV outcomes among adolescents and young people in Zambia.

## Background

The global HIV/AIDS epidemic remains a major public health concern, with adolescents and young people carrying a disproportionate burden, particularly in sub-Saharan Africa [1]. In 2023, an estimated 1.9 million adolescent girls and young women (15–24 years) were living with HIV, compared with 1.2 million adolescent boys and young men (15–24 years). Notably, 44% of all new HIV infections were among women and girls in 2023, with approximately 3,100 new cases occurring weekly in sub-Saharan Africa [2]. This highlights the vulnerability of this age group and the urgency of strengthening tailored interventions.

Adolescents and young people living with HIV (AYPLHIV) face unique developmental, psychosocial, and structural challenges that hinder their ability to access and adhere to HIV treatment and prevention services [3]. Among the populations most affected, adolescents represent a particularly vulnerable group. In Zambia, HIV prevalence among youths aged 15–24 years is estimated at 2.3% [4]. Stigma, limited confidentiality, and inadequate youth-friendly services remain major barriers. Addressing these challenges requires differentiated care approaches that adapt HIV services to the needs of specific populations, including self-care strategies [5].

Self-care, defined by the World Health Organization (WHO) as the ability of individuals, families, and communities to promote and maintain health, prevent disease, and cope with illness with or without the support of a healthcare provider, encompasses hygiene, nutrition, lifestyle, and socio-economic factors [6]. In HIV care, self-care services include ART adherence support, symptom self-monitoring, mental health self-care, and sexual and reproductive health practices [6]. These strategies support autonomy, resilience, and improved health outcomes during the transition to adulthood [7]. However, uptake of self-care services remains suboptimal due to stigma, limited awareness, socio-cultural norms, and service delivery gaps [6,8].

Evidence from low- and middle-income countries, particularly in sub-Saharan Africa, indicates that uptake of self-care services among AYPLHIV remains inconsistent and generally suboptimal. Although self-care interventions have been shown to improve treatment adherence and psychosocial well-being, their utilization is often limited by low awareness, persistent HIV-related stigma, and fear of unintended disclosure within households and communities [9,8,6]. Studies further highlight that adolescents are less likely to engage in self-care practices when youth-friendly services are unavailable, confidentiality is compromised, or healthcare providers display judgmental attitudes [10]. Socioeconomic constraints, including low levels of education and limited financial resources, further reduce adolescents’ capacity to independently access and sustain self-care behaviours.

Recent evidence suggests that self-care interventions can improve treatment adherence and quality of life among people living with HIV [11]. However, despite this growing body of global evidence, there remains a critical gap in context-specific knowledge regarding the uptake of self-care services among AYPLHIV in Zambia, where adolescent HIV remains a public health concern [12]. In particular, limited empirical data exist on the extent to which AYPLHIV utilize available self-care services and the factors that facilitate or hinder their uptake within routine HIV care settings. To address this gap, this study assessed the utilization of self-care services among adolescents and young people living with HIV in four health facilities in Kitwe District, focusing on levels of utilization and the individual, social, and health-system factors influencing uptake.

## Methods

### Research Design

This study employed a concurrent mixed-methods approach, combining quantitative cross-sectional design and qualitative case study design within a single study. The concurrent mixed methods design was used because it entails the researcher in gaining an in-depth understanding of the phenomena under the study and helps the researcher to understand the reality and how people interpret their experiences. This design was appropriate to allow a more comprehensive understanding of a research problem by drawing on the strengths of both quantitative and qualitative.

### Study Setting

The study was conducted between December 2024 and March 2025 in Kitwe District, located in the Copperbelt Province of Zambia. Kitwe is one of the major urban districts in the province and has a high burden of HIV, particularly among adolescents and young people. The district hosts several public health facilities providing comprehensive HIV care and treatment services, including adolescent- and youth-friendly health services.

Four health facilities were purposively selected for inclusion in the study due to their high caseloads of adolescents and young people living with HIV and the availability of youth-friendly spaces, which provide tailored services aimed at improving engagement, confidentiality, and retention in care among young clients. These characteristics made the selected facilities suitable for assessing the use of self-care services among AYPLHIV.

### Study Population

The study population comprised adolescents and young people aged 15–24 years living with HIV (AYPLHIV) who were receiving HIV care at the selected health facilities during the study period. In addition, adolescent health focal point persons working at these facilities were included to provide contextual insights into the provision and promotion of self-care services for AYPLHIV.

### Eligibility Criteria

Eligible participants were adolescents and young people aged 15–24 years living with HIV, registered for care and treatment at the selected health facilities, present during the data collection period, and willing to provide informed consent or assent, with parental or guardian consent obtained where required. Adolescents and young people who were severely ill, outside the eligible age range, or who declined participation were excluded from the study.

### Sampling techniques

For the quantitative component, a complete enumeration approach was employed, whereby all eligible adolescents and young people living with HIV (AYPLHIV) receiving care at the four purposively selected health facilities (Kawama, Luangwa, Ndeke, and Chimwemwe Health Facilities) in Kitwe District were targeted for inclusion. Participants were identified from facility HIV clinic registers and appointment lists for adolescents and young people receiving antiretroviral therapy.

Recruitment was conducted on-site at the health facilities during routine clinic days within the data collection period. Adolescents and young people who presented for HIV care were approached by trained research researcher, screened against the eligibility criteria, and invited to participate in the study. Those who met the inclusion criteria and provided informed consent or assent were enrolled. The selected health facilities functioned as natural clusters, and the enumeration of all eligible clients within these clusters minimized selection bias and enhanced statistical power by capturing the full variability of the target population.

### Data collection tools

Semi-structured questionnaires were used to collect detailed quantitative data on the uptake of self-care services among adolescents and young people living with HIV. The questionnaires were administered through face-to-face interviews conducted by trained research assistants to ensure clarity of questions, accommodate varying literacy levels, and enhance completeness of responses. Data was collected using a paper-based questionnaire, which was completed during routine clinic visits at the selected health facilities. The use of a face-to-face, paper-based approach allowed for immediate clarification of questions and minimized missing data. Completed questionnaires were reviewed daily for completeness before data entry into excel and Stata 17 for analysis.

### Dependent and individual level independent variables

In this study, the main outcome variable is the use of self-care services, measured by asking respondents whether they have ever accessed such services. Several independent variables are considered to help explain this outcome. Demographic characteristics include the age of adolescents and young people living with HIV, measured in exact years between 15 and 24; gender, categorized as male or female, the level of education, classified as no formal education, primary, secondary, or tertiary. Experiences of HIV stigma, also classified as low, moderate, or high; and the level of knowledge about HIV, determined by whether respondents demonstrate adequate understanding of the condition.

Accessibility and support factors include the availability of self-care services in the community, described as low, medium, or high, and the degree of family and peer support, also rated on the same scale. Behavioral factors are assessed through frequency of clinic visits categorized as rare, occasional, or regular, and the extent of peer influence, described as none, moderate, or high. Finally, organizational factors include the level of healthcare provider support, classified as low, moderate, or high. Together, these variables provide a comprehensive framework for examining the determinants of self-care service use among adolescents and young people living with HIV (see Table 1 below).

**Table 1:**
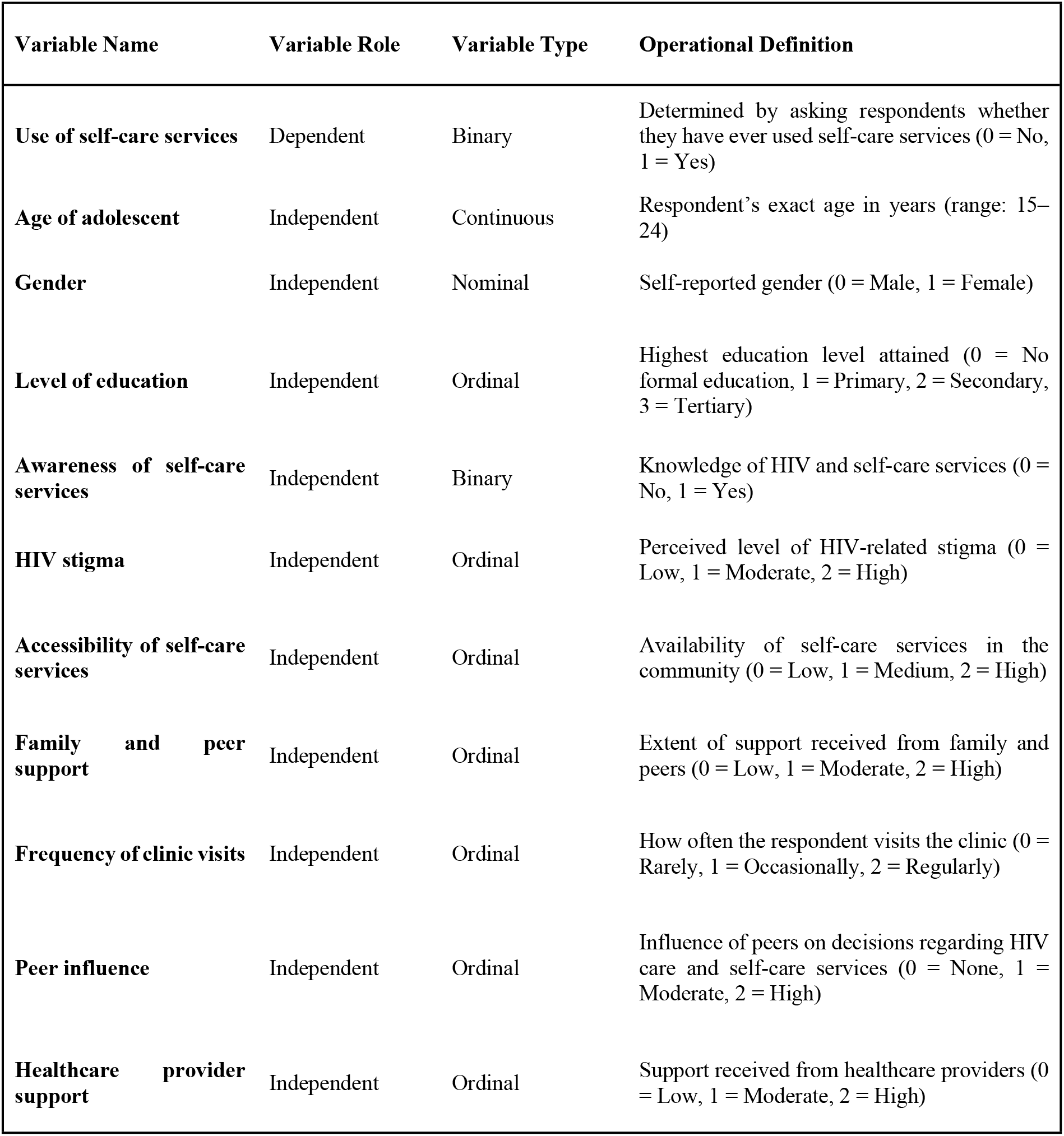
List of variables.

### Data Management and Analysis

Data from completed questionnaires were entered and analyzed using STATA version 17 for Windows. Prior to analysis, questionnaires were reviewed daily for completeness and consistency by the research team. Data entry was performed using a predefined coding framework, and double-checking of entries was conducted to minimize transcription errors. Range and consistency checks were applied to identify outliers, missing values, and illogical responses, which were resolved through verification with the original questionnaires where possible.

Descriptive statistics summarized participants’ demographic characteristics, with frequencies and percentages computed for categorical variables such as age group, gender, education level, and awareness of self-care services. Continuous variables were assessed for normality using the Shapiro-Wilk test. Associations between categorical variables and the outcome variable, “Use of Self-Care Services” (binary: Yes/No), were initially evaluated using Chi-square or Fisher’s exact tests, as appropriate. A multivariable logistic regression model was then fitted to identify independent factors associated with self-care service utilization, with model diagnostics including the Akaike Information Criterion (AIC), Bayesian Information Criterion (BIC), and checks for multicollinearity among predictors. Statistical significance was assumed at an alpha level of 0.05.

### Ethical Clearance

Ethical clearance for the study was obtained from the University of Zambia Biomedical Research Ethics Committee (UNZABREC), **reference number 6042-2024**, along with written approvals from other relevant authorities prior to commencing the study. The study adhered to key ethical principles, including respect for participants’ privacy, confidentiality, and voluntary participation. Participation was entirely voluntary, with participants free to decline or withdraw at any stage without any consequences for the study outcomes. Data collection procedures, including interviews and questionnaires (10–15 minutes each) and focus group discussions (20–30 minutes), were designed to minimize burden on participants. Informed consent, anonymity, and confidentiality were strictly maintained. Furthermore, identification numbers were assigned to questionnaires to ensure participant confidentiality.

## Results

### Background characteristics of participants

The study’s findings reveal several notable patterns among adolescents and young people. The results in table 2 show the socio-demographic characteristics of the four hundred and eight-five (n=485) respondents revealing a balanced representation across key variables. The majority (54%) were aged 15-19 years, while 46% were 20-24 years, indicating that younger adolescents form a slightly larger proportion of the study population. Gender distribution was nearly equal, with 49% male and 50% female, ensuring that the findings reflect the experiences of both genders. In terms of education, 16% had no formal education, 15% had primary education, 23% had secondary education, and 45% had tertiary education (see Table 2 below).

**Table 2:** Socio-demographic demographic characteristics of the respondents, (n = 485)

| Variable | Frequency | Percentage |
| --- | --- | --- |
| **Age** |  |  |
| 15-19 Years | 260 | 53.61 |
| 20-24 Years | 225 | 46.39 |
| **Gender** |  |  |
| Male | 241 | 49.49 |
| Female | 244 | 50.31 |
| **Education Level** |  |  |
| No education | 78 | 16.08 |
| Primary | 74 | 15.26 |
| Secondary | 113 | 23.30 |
| Tertiary | 220 | 45.36 |

### Awareness of Self-Care Services among Adolescents and Young People

The pie chart illustrates the awareness of self-care services among adolescents and young people living with HIV/AIDS. Most respondents (56%, n=274) reported being not aware of self-care services, while 44% (n=211) indicated being aware of the services at the facility (see Fig 1).

**Figure 1:**
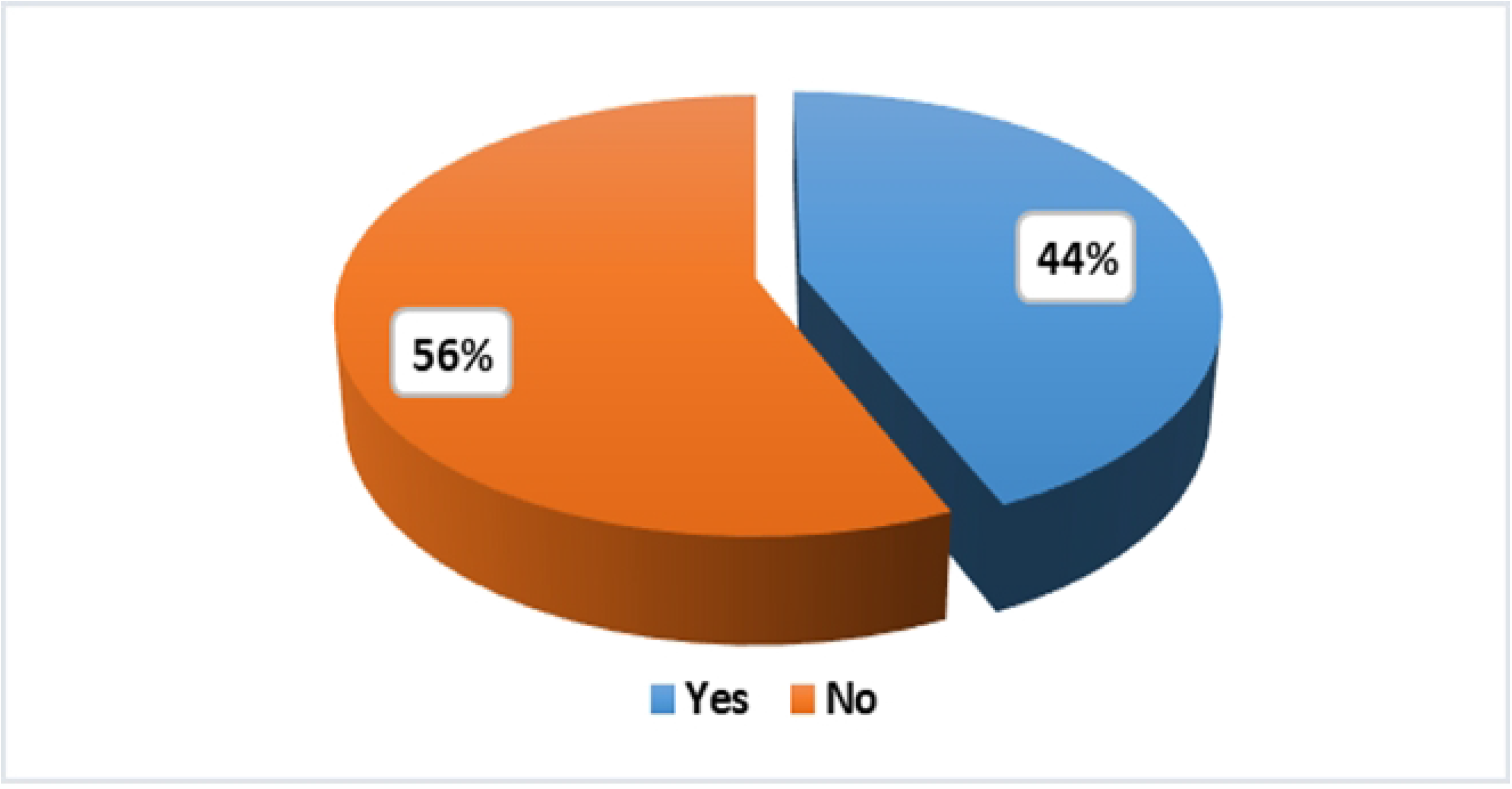
Awareness of Self-Care Services among Adolescents and Young People.

### Use of self-care services among adolescents and young people

Out of 485 respondents, 280 individuals (58%) reported that they have used self-care services before, however, a significant proportion (42%, n=205 respondents) have never used these services, highlighting a potential gap in service use (see Fig 2).

**Figure 2:**
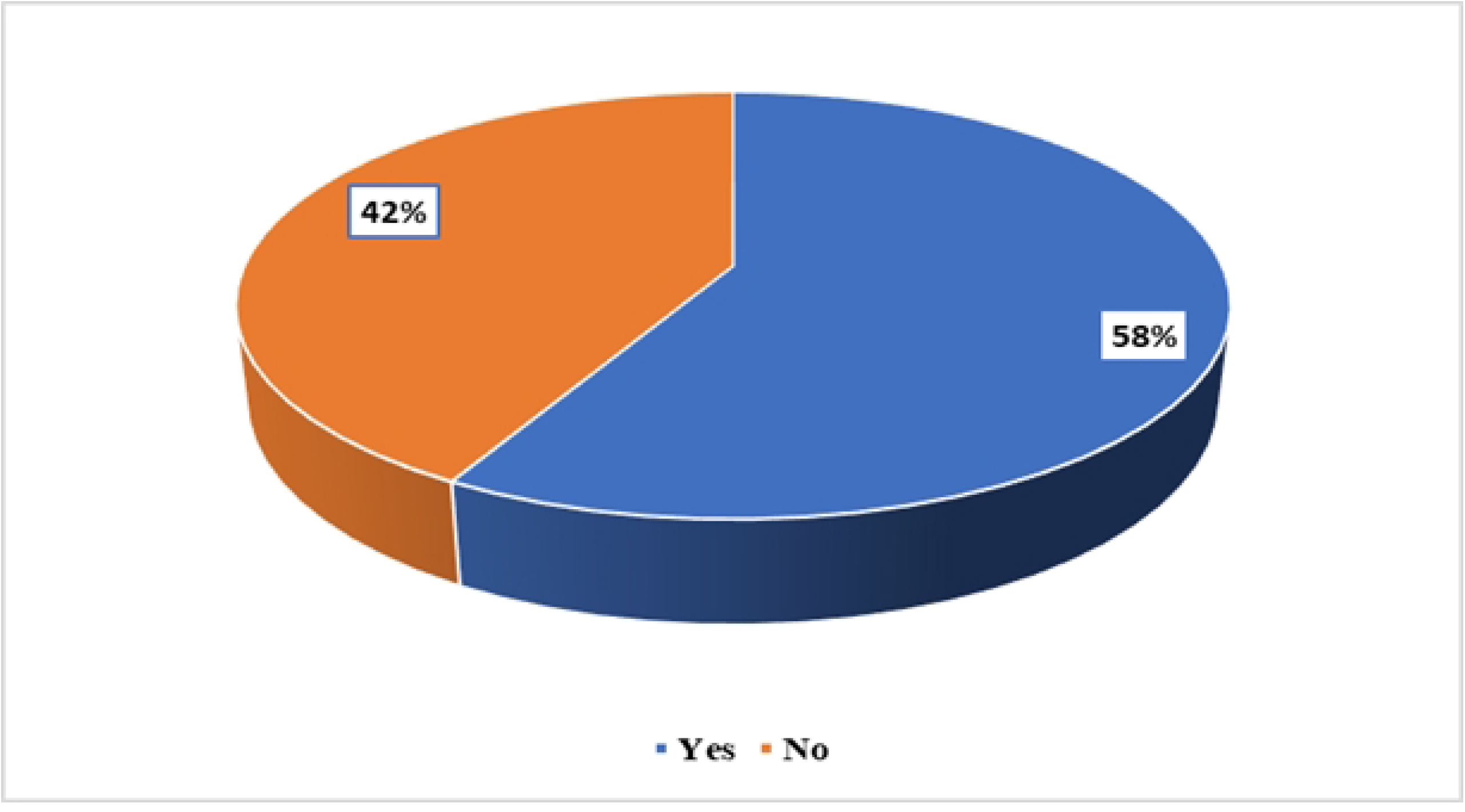
Adolescents and young people ever used self-care services.

### Analysis of factors associated with the use of self-care services

The analysis in table 3 revealed that age was not significantly associated with the use of self-care services (p=0.869). Both adolescents aged 15–19 years (41.9%) and those aged 20–24 years (42.7%) reported similar usage levels. Gender showed a borderline association with self-care use (p=0.070), with a higher proportion of females (46.3%) utilizing services compared to males (38.2%). A strong association was observed between education level and the use of self-care services (p=0.013). Respondents with tertiary education reported the highest use (48.2%), while those with secondary education recorded the lowest (30.1%). Conversely, HIV knowledge confidence did not demonstrate any significant relationship (p=0.808), as uptake rates were consistent across levels of confidence. Similarly, receipt of HIV information (p=0.490) and awareness of self-care services (p=0.541) did not significantly suggest association.

**Table 3:** Variable distribution and association of adolescent and young people’s use of selfcare services.

| Variables | Used Self Care Services |  | P-Value |
| --- | --- | --- | --- |
|  | No | Yes |  |
| **Age** |  |  |  |
| 15-19 | 151 (58.08) | 109 (41.92) | 0.869 |
| 20-24 | 129 (57.33) | 96 (42.67) |  |
| **Gender** |  |  |  |
| Female | 131 (53.69) | 113 (46.31) | 0.07 |
| Male | 149 (61.83) | 92 (38.17) |  |
| **Education Level** |  |  |  |
| No Education | 42 (53.85) | 36 (46.15) | 0.013*** |
| Primary | 45 (60.81) | 29 (39.19) |  |
| Secondary | 79 (69.91) | 34 (30.09) |  |
| Tertiary | 114 (51.82) | 106 (48.18) |  |
| **HIV Knowledge Confidence** |  |  |  |
| Not Confident at All | 96 (56.47) | 74 (43.53) | 0.808 |
| Somewhat Confident | 93 (57.06) | 70 (42.94) |  |
| Very Confident | 91 (59.87) | 61 (40.13) |  |
| **Received HIV Information** |  |  |  |
| No | 140 (56.22) | 109 (43.78) | 0.49 |
| Yes | 140 (59.32) | 96 (40.68) |  |
| **Awareness of Selfcare services** |  |  |  |
| No | 177(58.80) | 124 (41.20) | 0.541 |
| Yes | 103 (55.98) | 81 (44.02) |  |
| **Support from Health Providers** |  |  |  |
| Low | 86 (57.72) | 63 (42.28) | 0.488 |
| Moderate | 91 (54.49) | 76 (45.51) |  |
| High | 103 (60.95) | 66 (39.05) |  |
| **Peer Influence** |  |  |  |
| No peer influence | 113 (58.25) | 81 (41.75) | 0.497 |
| Moderate | 91 (60.67) | 59 (39.33) |  |
| High | 79 (53.90) | 65 (46.10) |  |
| **HIV Stigma** |  |  |  |
| Low | 86 (49.71) | 87 (50.29) | 0.024*** |
| Moderate | 57 (64.77) | 31 (35.23) |  |
| High | 137 (61.16) | 87 (38.84) |  |
| Family and peer support |  |  |  |
| Low | 90 (60.40) | 59 (39.60) | 0.261 |
| Moderate | 135 (59.21) | 93 (40.79) |  |
| High | 55 (50.93) | 53 (49.07) |  |
| Accessibility of Self-Care Services |  |  |  |
| Not Accessible | 81 (64.80) | 44 (35.20) | 0.041** |
| Partially Accessible | 83 (61.03) | 53 (38.97) |  |
| Accessible | 116 (51.79) | 108 (48.21) |  |

Statistical significance is denoted as \*\*\* $p < 0.01$ , \*\* $p < 0.05$ , and $p \geq 0.05$ considered not statistically significant. All tests were conducted using the Chi-square tests

In terms of reasons for non-use, the association was marginal (p=0.079). Notably, respondents citing “no need” for services reported higher usage (52.6%) compared to those mentioning access issues (35.4%), suggesting that structural barriers may still limit uptake. Support from health providers also showed no significant association, although moderate support was linked with higher use (45.5%). Likewise, peer influence was not significantly associated (p=0.497), even though respondents experiencing high peer influence had slightly greater uptake (46.1%).

Importantly, HIV stigma demonstrated a significant association with self-care service use (p=0.024). Respondents experiencing low stigma reported the highest usage (50.3%), while those with moderate stigma had the lowest (35.2%). Finally, accessibility of self-care services was also significantly associated (p=0.041). Those reporting that services were accessible had higher usage (48.2%) compared to those who indicated services were not accessible (35.2%), see Table 3.

### Factors Associated with the use of self-services among adolescents and young people

The analysis revealed that age was not a significant predictor of self-care service use. Respondents aged 20–24 years were almost equally likely to use self-care services as those aged 15–19 years, with both the unadjusted and adjusted models showing odds ratios close to one and p-values greater than 0.8. Similarly, gender did not significantly influence self-care service use, although the adjusted results suggested that males were somewhat less likely to use such services compared to females (OR = 0.72, 95% CI: 0.50–1.05, p = 0.085).

Education level, however, showed some association. Participants with secondary education were less likely to use self-care services compared to those with no education, and this finding was statistically significant in the adjusted model (OR = 0.54, 95% CI: 0.29–1.00, p = 0.048). Primary and tertiary education, on the other hand, did not show significant association. Receiving information on HIV and being aware of self-care services were also not significantly associated with service use.

Accessibility of services emerged as a predictor. Respondents who reported that self-care services were accessible had significantly higher odds of utilizing them compared to those who considered them not accessible (adjusted OR = 1.67, 95% CI: 1.04–2.68, p = 0.031). Partial accessibility, however, was not associated with increased utilization. Clinic visit frequency showed no significant effect, as those who visited occasionally, rarely, or regularly did not differ in their likelihood of using self-care services. Similarly, the level of support from providers and peer influence were not statistically significant factors.

HIV stigma was another important determinant. Individuals who experienced moderate stigma were significantly less likely to use self-care services compared to those with low stigma (adjusted OR = 0.53, 95% CI: 0.30–0.91, p = 0.022). Likewise, high stigma was negatively associated with the use of selfcare services (adjusted OR = 0.63, 95% CI: 0.41–0.95, p = 0.029), see Table 4.

**Table 4:** Multivariable analysis of the factors associated with the use of self-care Services.

| Variable | Unadjusted |  |  | Adjusted |  |  |
| --- | --- | --- | --- | --- | --- | --- |
|  | OR | 95% CI | P-value | OR | 95% CI | P-value |
| **Age group** |  |  |  |  |  |  |
| 15-19 | Ref | - | - | Ref | - | - |
| 20-24 | 1.03 | (0.86, 1.48) | 0.869 | 1.00 | (0.69, 1.47) | 0.971 |
| **Gender** |  |  |  |  |  |  |
| Female | Ref | - | - | Ref | - | - |
| Male | 0.72 | (0.50, 1.03) | 0.07 | 0.72 | (0.50, 1.05) | 0.085 |
| **Education** |  |  |  |  |  |  |
| No level of education | Ref | - | - | Ref | - | - |
| Primary | 0.75 | (0.39, 1.43) | 0.386 | 0.77 | (0.39, 1.49) | 0.431 |
| Secondary | 0.50 | (0.28, 0.91) | 0.024 | 0.54 | (0.29, 1.00) | 0.048 |
| Tertiary | 1.08 | (0.65, 1.82) | 0.758 | 1.11 | (0.65, 1.90) | 0.713 |
| **Received information on HIV** |  |  |  |  |  |  |
| No | Ref | - | - | Ref | - | - |
| Yes | 0.88 | (0.61, 1.26) | 0.49 | 0.84 | (0.58, 1.24) | 0.399 |
| **Awareness of Self-Care Services** |  |  |  |  |  |  |
| No | Ref | - | - | Ref | - | - |
| Yes | 1.26 | (0.77, 1.63) | 0.541 | 1.03 | (0.70, 1.52) | 0.879 |
| **Accessibility of Self-Care Services** |  |  |  |  |  |  |
| Not Accessible | Ref | - | - | Ref | - | - |
| Partially Accessible | 1.18 | (0.71, 1.95) | 0.529 | 1.15 | (0.68, 1.99) | 0.546 |
| Accessible | 1.71 | (1.09, 2.69) | 0.019 | 1.67 | (1.04, 2.68) | 0.031*** |
| **Clinic Visit Frequency** |  |  |  |  |  |  |
| Occasionally visit | Ref | - | - | Ref | - | - |
| Rarely visit | 0.78 | (0.50, 1.21) | 0.263 | 0.76 | (0.48, 1.21) | 0.247 |
| Regularly visit | 0.88 | (0.57, 1.37) | 0.575 | 0.88 | (0.55, 1.40) | 0.596 |
| **Support from providers** |  |  |  |  |  |  |
| High | Ref | - | - | Ref | - | - |
| Moderate | 1.3 | (0.85, 2.01) | 0.231 | 1.11 | (0.85, 2.11) | 0.208 |
| Low | 1.14 | (0.73, 1.79) | 0.559 | 1.34 | (0.70, 1.78) | 0.657 |
| **Peer Influence** |  |  |  |  |  |  |
| Low | Ref | - | - | Ref |  |  |
| Moderate | 0.9 | (0.59, 1.40) | 0.651 | 0.95 | (0.70, 1.78) | 0.644 |
| High | 1.19 | (0.77, 1.85) |  | 1.2 | (0.86, 2.13) | 0.197 |
| **HIV Stigma** |  |  |  |  |  |  |
| Low | Ref |  |  | Ref |  |  |
| Moderate | 0.53 | (0.32, 0.91) | 0.022 | 0.53 | (0.30, 0.91) | 0.022*** |
| High | 0.63 | (0.42, 0.94) | 0.023 | 0.63 | (0.41, 0.95) | 0.029*** |

\*\*\*P<0.05

## Discussion

The findings of this study provide important insights into the socio-demographic characteristics and factors influencing the utilization of self-care services among adolescents and young people living with HIV. This study examined the level of utilization of self-care services and factors influencing uptake among AYPLHIV in four health facilities in Kitwe District. The study found no statistically significant association between age or gender and the use of self-care services, although females demonstrated slightly higher uptake than males. This finding is consistent with studies conducted in sub-Saharan Africa, which report minimal age and gender differences in self-care or HIV service utilization once adolescents are engaged in care [13,7]. The absence of strong demographic effects suggests that individual characteristics alone may be insufficient to explain self-care behaviour among AYPLHIV, highlighting the importance of contextual and service-related factors.

Educational attainment emerged as a significant predictor of self-care utilization, with adolescents and young people who had secondary education being less likely to use self-care services compared to those with no formal education. This counterintuitive finding contrasts with existing literature that generally associates higher education with improved health literacy and service utilization [11]. However, similar patterns have been reported among school-going adolescents living with HIV, where competing academic demands, fear of stigma within school settings, and concerns about disclosure limit engagement with HIV-related services [12,14]. This underscores the need for school-sensitive and adolescent-friendly self-care interventions.

Accessibility of self-care services was a strong facilitator of utilization in this study. Adolescents who perceived services as accessible were significantly more likely to use them. This finding aligns with evidence from sub-Saharan Africa demonstrating that physical proximity, confidentiality, flexible clinic hours, and youth-friendly environments are critical determinants of adolescent HIV and sexual and reproductive health service uptake [15,16]. The WHO similarly emphasizes that adolescent-responsive health systems characterized by respectful provider attitudes and convenient service delivery are essential for improving engagement in HIV prevention, treatment, and self-care [13]. When services are perceived as accessible and non-judgmental, adolescents are more likely to seek care, adhere to treatment, and adopt self-care practices [2].

HIV-related stigma emerged as a major barrier to self-care utilization, with adolescents experiencing moderate to high levels of stigma being less likely to engage in self-care practices. This finding is consistent with extensive evidence demonstrating that stigma, fear of disclosure, and anticipated discrimination undermine adolescents’ engagement with HIV-related services [17]. Intersectional stigma related to age, HIV status, and social identity further constrains access to care at individual, interpersonal, and structural levels [18]. In Zambia, qualitative studies indicate that fear of unintended disclosure and perceived judgment from healthcare workers and community members discourage adolescents from seeking care and engaging in self-care practices [19,14]. Similar findings from other African settings show that stigmatizing provider behaviours and fear of negative peer reactions reduce adolescents’ confidence in accessing HIV services [20,21].

### Implications for policy and practice

The findings suggest that improving uptake of self-care services among AYPLHIV requires holistic strategies that combine youth-friendly service delivery, stigma reduction, targeted education, and supportive environments. Policies should prioritize confidentiality, accessibility, and adolescent-sensitive care. National HIV guidelines should further integrate self-care interventions such as HIV self-testing, digital adherence support, and community-based ART refills. Stigma reduction initiatives involving communities, families, and peer networks are essential to creating enabling environments for adolescents.

Finally, policy frameworks must commit adequate financial resources, strengthen multi-sectoral partnerships, and institutionalize robust monitoring and evaluation systems to ensure sustainability. However, Zambia’s health sector remains constrained by donor dependence and limited domestic financing [22]. Addressing these structural and funding barriers is critical to ensuring that self-care services are not only available but actively utilized, thereby improving autonomy, health outcomes, and quality of life among adolescents and young people living with HIV.

### Strengths and limitations of the study

This study provides important empirical evidence on the utilization of self-care services among adolescents and young people living with HIV in Zambia, a context where such evidence remains limited. The inclusion of four health facilities and a relatively large sample size strengthened the representativeness of the findings within Kitwe District. Complete enumeration of eligible adolescents minimized selection bias, while the use of multivariable logistic regression enhanced analytical rigor by adjusting for potential confounders. In addition, incorporating perspectives from healthcare providers enriched the analysis by identifying health-system–level barriers and opportunities relevant to policy and practice.

Nevertheless, several limitations should be considered. The cross-sectional study design precludes causal inference between identified factors and self-care service utilization. Reliance on self-reported data may have introduced social desirability and recall biases, particularly given the sensitive nature of HIV-related behaviours. Furthermore, the restriction of the study to four health facilities within a single district may limit the generalizability of findings to other regions of Zambia with different socio-cultural and health-system contexts. Finally, resource and time constraints prevented longitudinal follow-up, which could have provided deeper insight into changes in self-care utilization patterns over time.

## Conclusion

This study examined the utilization of self-care services and associated factors among adolescents and young people living with HIV attending selected health facilities in Kitwe District, Zambia. The findings indicate that, despite growing recognition of self-care as a critical component of HIV management, uptake among adolescents and young people remains suboptimal. Although some participants reported engaging in self-care practices, substantial gaps in awareness, accessibility, and supportive environments continue to constrain effective utilization. Education level, service accessibility, and HIV-related stigma emerged as key determinants of self-care service use, while age and gender were not significantly associated with uptake. These results underscore the need to address structural and psychosocial barriers rather than focusing solely on demographic characteristics. In particular, limited access to services and persistent stigma within communities and health facilities undermine adolescents’ capacity to manage their health independently and fully benefit from available self-care interventions.

## Data Availability

The dataset has been uploaded for reference

## Acknowledgments

We extend our sincere gratitude to the adolescents and young people, their parents and guardians, as well as the healthcare providers and the Adolescent Health Focal Point Person (ADH-FPP) whose participation and support made this study possible.

## Author Contributions

Conceptualization: Elvis Dala Makukula Data curation: Elvis Dala Makukula Formal analysis: Elvis Dala Makukula

Methodology: Elvis Dala Makukula, Samson Shumba, Chishimba Nakamba Mulambia, Choolwe Jacobs

Software: Elvis Dala Makukula

Visualization: Samson Shumba, Chishimba Nakamba Mulambia Writing – original draft: Elvis Dala Makukula

Writing – review and editing: Elvis Dala Makukula, Samson Shumba, Chishimba Nakamba Mulambia, Choolwe Jacobs

Supervisor: Choolwe Jacobs

